# Experiences of breastfeeding peer support: views from current, past and trainee breastfeeding peer support volunteers in Wales, UK

**DOI:** 10.1101/2024.05.15.24307400

**Authors:** H Jones, R Davies, F Williams, R Evans, R Bath, R Embling, S Brophy

**Affiliations:** National Centre for Population Health and Wellbeing Research; Public Health Wales

## Abstract

**Background:** Breastfeeding peer support involves mothers with personal experience of breastfeeding providing support, information, and encouragement to other breastfeeding mothers. The Unicef UK Baby Friendly Initiative standards require that additional support for new mothers, such as peer support, is provided and that services work together to ensure that mothers can get additional help as needed. As work begins on a national framework for feeding support, we wanted to explore the current activity of breastfeeding supporters in Wales, UK. We aimed to explore the experiences of current, past and trainee breastfeeding peer support volunteers to understand what they need from a refreshed All Wales Breastfeeding Action Plan and how they could benefit.

**Methods:** Current, past and trainee breastfeeding peer support volunteers were invited by Public Health Wales to complete an online survey about their experiences of providing breastfeeding peer support including both open and closed questions. Codebook thematic analysis was used to generate key themes arising from qualitative data.

**Results:** 81 volunteers completed the online survey between 21/12/23 and 22/1/24. Their experiences were described as ’rewarding’ (50% (44/81)) and ‘empowering’ (12%, 10/81), but also ’frustrating’ (19%, 15/81) and ’challenging’ (8%, 7/81). Peer supporters supported basic positioning and latching, challenges with milk supply, frequency of feeds, weaning, and poor weight gain. Three key themes were developed from the open-ended question ‘why did you decide to undertake peer support training?’ (1) Paying it forward (2) Limited peer support in local areas and (3) The need for more support. Funding was noted as a barrier to continuing to provide breastfeeding peer support.

**Conclusions:** Breastfeeding peer support was associated with benefits for the peer supporters themselves and recognised as beneficial for the mothers they helped. Investment in training future breastfeeding peer supporters is essential to ensure the continued success of these important initiatives.

## Introduction

Peer support refers to the provision of assistance, guidance and encouragement by individuals who share similar experiences or backgrounds [1]. In the context of breastfeeding, peer support involves mothers with experience in breastfeeding that have undertaken the relevant training in order to provide support, information, and encouragement to other breastfeeding mothers [2]. Breastfeeding peer support is part of the standard pathway, linking mothers who want to breastfeed to others who have personal experience and some training. The Unicef UK Baby Friendly Initiative standards require that additional support for new mothers such as peer support is provided and that services work together to ensure that mothers can get additional help as needed [3].

Previous research has facilitated the understanding of breastfeeding peer support including the experiences and perceptions of mothers who received breastfeeding peer support [4]. Experiences of peer support ranged from authentic presence to disconnected encounters [4]. One study evaluated the effectiveness of a peer support initiative, which trained peer supporters who then set up a support group, in an area of social and economic deprivation in England, UK [5]. This research helped areas that tend to face more barriers and challenges and have fewer breastfeeding role models, by increasing the number of women who initiate and continue to breastfeed [5]. Breastfeeding peer support is a valuable and evidence-based intervention that has a positive impact on mothers achieving their breastfeeding goals and enhancing their wellbeing [6]. These findings demonstrate that peer support volunteers can offer optimal and tailored support, while also improving their own skills and motivation in the process.

Previously a systematic review was conducted to examine the effectiveness of community-based peer support for breastfeeding mothers [7]. The findings highlighted that community-based peer support for mothers is effective in increasing the duration of exclusive breastfeeding, particularly for infants aged 3–6 months in low and middle-income countries. Such support also encourages mothers to initiate breastfeeding early [7]. Research has indicated that breastfed mothers are more likely to breastfeed their children [8]. Therefore, peer support is important to support breastfeeding and close potential inequalities.

A survey of all UK infant feeding coordinators who were part of UK-based National Infant Feeding Networks, covering 177 National Health Service (NHS) organisations, recorded that there was a lack of information about how, when and where breastfeeding peer support was provided in the UK [9]. They found that the provision of training and peer supporter roles varied significantly between services. One third of respondents felt that breastfeeding peer support services were not well integrated with the NHS and one quarter of respondents stated that breastfeeding peer support was not accessed by mothers from poorer social backgrounds [9].

Previous research has explored breastfeeding peer supporters’ experiences of volunteering within hospitals [10]. Volunteers were motivated by wanting to help, their future career and having social contact. Positive experiences of voluntary activity made peer supporters feel valued.

However, there were areas within this study where the peer support service needed more investment. This included the organisational support for the volunteer system and calls for improved planning were also highlighted as integral to ensure the sustainability of the service [10].

A study that explored the use of peer support in England focused on the views of mothers, midwives and peer supporters and the effects on their breastfeeding experiences. Their findings suggested that a targeted peer support service was associated with small non-significant increases in breastfeeding rates. They concluded that peer support was associated with psycho- social benefits for mothers, health professionals and peer supporters [11].

In addition to supporting breastfeeding rates, peer support initiatives can more broadly support the wellbeing of breastfeeding mothers. Recent research examined qualitative evidence of women’s, peer supporters’ and healthcare professionals’ views and experiences of breastfeeding peer support [12]. They reported positive characteristics, approaches and benefits of peer support and supporters. The findings represented that breastfeeding peer support increased women’s self-esteem and confidence in breastfeeding while reducing social isolation. They offer practical advice, encouragement, reassurance and emotional support based on their own experiences. Peer supporters valued the experience, which gave them a sense of purpose and confidence, and felt good about helping the women they supported [12]. Women appreciated peer supporters who were caring, spent time with them, shared experiences, provided realistic information along with practical and emotional support [12].

### All Wales Breastfeeding Action Plan

The aim of the All Wales Breastfeeding Action Plan (AWBAP) is that more babies in Wales will be breastfed and for longer with a reduction in inequalities [13]. The AWBAP recommends that a coordinated support model should be developed across Wales to ensure appropriate care for breastfeeding dyads [13]. This model would incorporate peer support with all of its potential benefits to mothers and to volunteers. Furthermore, the AWBAP aims to take specific action to develop national guidance on peer support where the voice of peer supporters themselves inform this guidance. However, there is a need to understand the current extent of peer support in Wales, in order to inform the development of national policy.

### Aims and Hypotheses

As work begins on a national framework for feeding support, we wanted to explore the current activity of breastfeeding supporters across Wales using an online survey with both quantitative and qualitative responses. Results from this study will be used to understand the current extent of peer support in Wales and inform the development of national policy or guidance on breastfeeding peer support and recognition of the vital service peer supporters provide. The purpose of this work was to explore the experiences of current, past and trainee breastfeeding peer support volunteers in Wales, UK.

## Methods

### Survey for Peer support volunteers

Current, past and trainee breastfeeding peer support volunteers in Wales were invited to complete an online survey via social media advertising/email. Consent was implied through individuals’ decision to participate in the voluntary anonymous survey after being provided with information about the survey’s purpose and how findings would be used. The survey took approximately 10 minutes to complete. Closed questions were used to ascertain information about participants’ volunteer experience and training status. Open questions were included to gain an understanding of why they chose to participate in peer support and to provide rich details of their experience. The survey questions can be found in the supplementary information.

Quantitative survey data were summarised using descriptive statistics as well as identifying frequencies of concepts in survey responses. Codebook thematic analysis [14] was used to generate themes from an open-ended question on the survey: ‘Why did you decide to undertake peer support training?’ Thematic analysis identifies and describes patterns across data. ‘Codebook’ approaches use a structured coding framework to develop and document the analysis [14,15]. Analysis involved six phases 1) data familiarisation and writing familiarisation notes 2) systematic data coding 3) generating initial themes from coded and collated data 4) developing and reviewing themes 5) refining, defining, and naming themes and 6) writing the report. All data were independently analysed by HJ and FW, who then discussed their findings. This was to ensure that important concepts within the data were not missed, and to achieve a richer understanding of the data through multiple perspectives.

## Results

The survey received 81 responses between 21/12/23 and 22/1/24. Of those, n=45 were currently active as a peer support volunteer within a breastfeeding organisation (a group of trained volunteers who support breastfeeding mothers e.g. Association of Breastfeeding Mothers (ABM)), while n=14 were active with the local NHS. Of these volunteers, n=13 were previously active with a breastfeeding organisation, n=8 were previously active in the NHS and n=13 were trained or in training but had not yet been active (see Figure 1). Overall, 73% of respondents were currently active peer supporters (n=59) either with a breastfeeding organisation and/or the NHS. The majority of respondents have been volunteering as peer supporters for more than two years (see Figure 2). The majority spent about 2 hours a week volunteering (29.6%, 24/81) with (25.9%, 21/81) spending more than 2 hours and some also reported spending additional time sharing support and answering questions via online services or social media. It must be noted that there is no up to date record or register of the total number of peer supporters in Wales.

**Figure 1.**
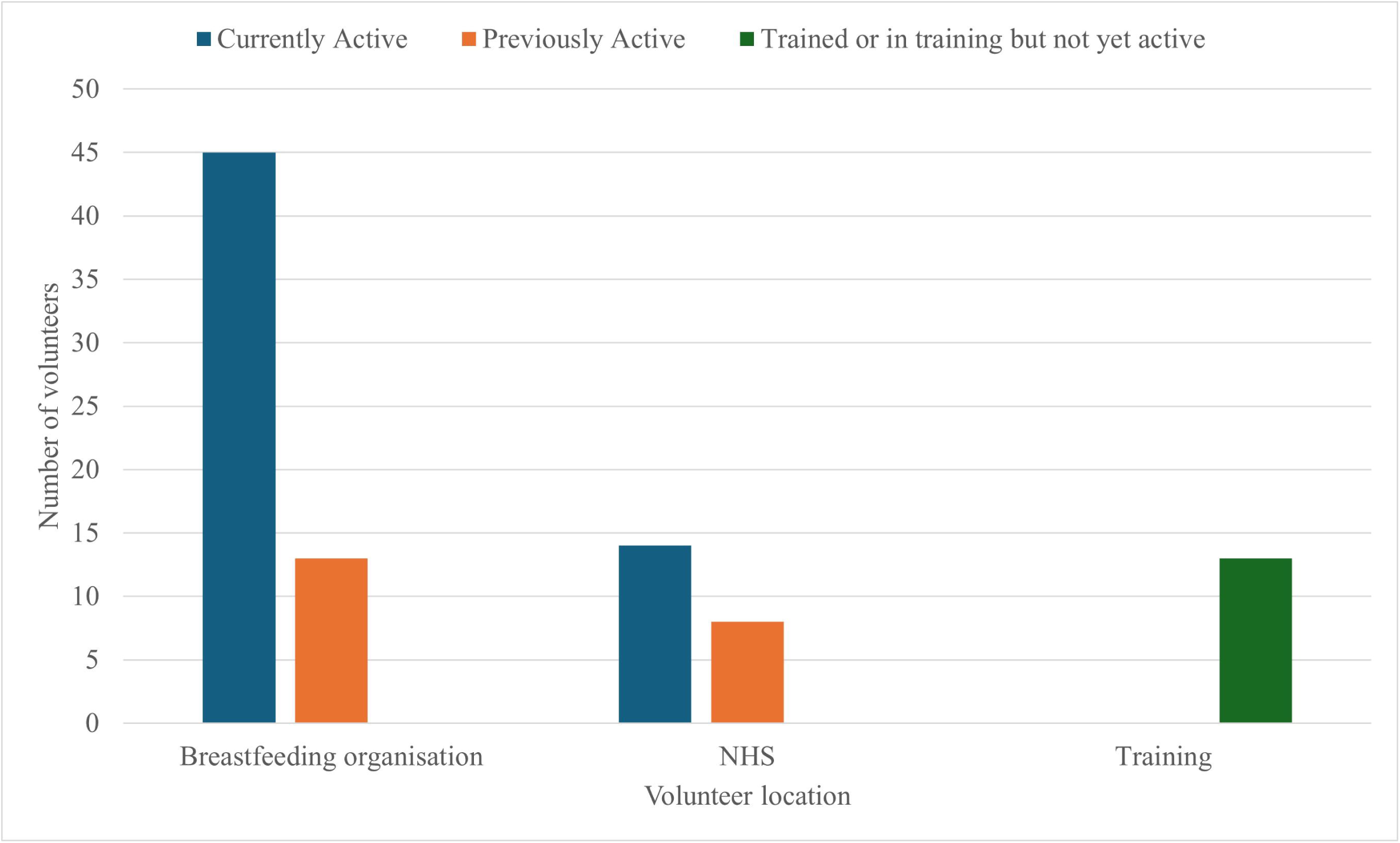
Number of survey respondents that were currently active, previously active, or in training and where they volunteered.

**Figure 2.**
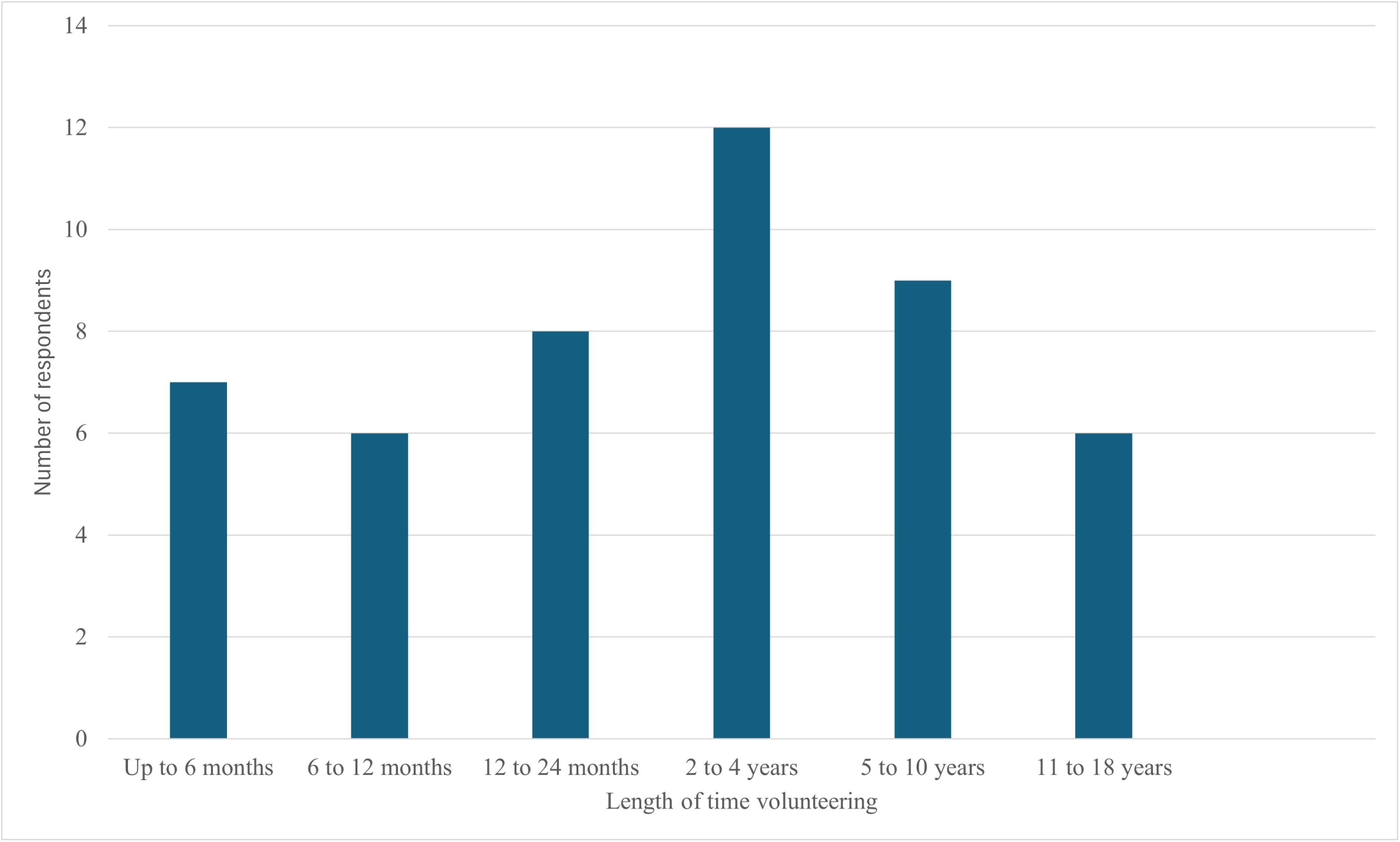
Length of time volunteers provided peer support.

## Experiences

When asked to describe the experience of breastfeeding peer support the majority felt it was ’rewarding’ (50% (44/81)) and ‘empowering’ (12%, 10/81), but also ’frustrating’ (19%, 15/81) and ’challenging’ (8%, 7/81). However, they also raised concerns about funding (19.8% of respondents). Volunteers highlighted that they felt they were being used to fill funding gaps within NHS services (n=9). Comments included thoughts that “peer supporters should be in addition to NHS support not instead of it” and “there should be far more funding and provisions to install peer supporters within hospitals”.

When asked about what were the most common issues for which support was needed, (18.5%, 15/81) said basic positioning and challenges with milk supply, whilst (19.7%, 16/81) commented about tongue tie and poor weight gain. Frequency of feeds (n=<5) and weaning (n=<10).

## Motivation

An open ended question on the survey asked why they decided to undertake peer support training. Three key themes were developed from the qualitative data: (1) Paying it forward.

Many respondents reported that they had received support themselves and wanted to provide the same for others, “To pay it forward” and “I received peer support so was keen to help others in turn”. (2) Limited support locally. “I wanted to help other mums in the area as there were limited peers in my area”, “There wasn’t a group local to me” and “Breastfeeding support in West Wales is very minimal”. 3) Lack of information. “there is a dire need for information and support for breastfeeding” and “Because of the lack of support I had after my first baby.” The viewpoints shared have highlighted the important role that peer supporters play in helping women to have positive breastfeeding journeys “it was peer support which helped me with my own breastfeeding journey struggles when NHS support was woefully lacking.” “ I was passionate about supporting mothers, especially those who had meant their struggles and lack of support had led them to stopping.”

## National guidance

Among the 81 respondents, n=56 felt that national guidance for infant feeding teams on peer support was essential. Additionally, n=21 respondents acknowledged that national guidance would be useful. The consensus was that national guidance would be beneficial to infant feeding teams, enhancing their effectiveness and support networks: n=41 of respondents said that it was essential to have national peer support training or a qualification, with a further n=38 saying it would be useful or nice to have. There was a strong inclination towards the potential benefits and importance of implementing a standardised national qualification or training programme.

## Funding

Peer supporters expressed frustrations at the personal financial impact that volunteering has had on them, with one respondent stating that they were ‘out of pocket’ paying for membership fees and refreshments to help run the groups. Another respondent expressed the need to “pay to run our peer supporter course and then fundraising and organising funding to train following cohorts”. Nearly half of the peer supporters were self-funded (48.1%, 39/81) with (18.5%, 15/81) being NHS funded and (29.6%, 24/81) charity project funded. This highlights that a relatively high percentage of peer supporters are self-funding their training costs and expenses needed to undertake their volunteering work.

Of the 17 women who were no longer peer supporters they reported this was because they were too busy (n=<10), returned to work (n=<10), started midwifery/nursing training or another breastfeeding related role (n=<5).

## Discussion

Findings demonstrate that the experiences of current, past and trainee breastfeeding peer support volunteers in Wales is both rewarding, empowering and frustrating. Breastfeeding peer support volunteers described the positive experiences of receiving breastfeeding peer support themselves and wanting to help other breastfeeding mothers have the same experience. The volunteers highlighted that more peer support and professional infant feeding support needs to be available in Wales and further training opportunities are needed.

Ensuring the quality and consistency of support provided by peer supporters can be challenging. While training is typically provided, there may be variations in the quality of training and supervision across different programs. Inconsistent support or misinformation from poorly trained peer supporters could undermine the effectiveness of peer support programs in Wales. This provides further support for there to be national standardisation of training, which is also favoured by respondents to the survey. Sustaining breastfeeding peer support programs in Wales may require ongoing investment and commitment from policymakers and healthcare providers. The financial impact that is at times put on some peer supporters to undertake their voluntary work is a concern that should be addressed. Without adequate resources and support, there is a risk of volunteer burnout, high turnover rates among peer supporters, or the discontinuation of programs, which could limit their long-term impact.

## Strengths and weaknesses

Breastfeeding peer support programs have previously been associated with increased breastfeeding initiation and continuation rates [4–7]. By providing practical guidance, emotional support, and sharing personal experiences, peer supporters can positively influence mothers’ breastfeeding decisions and practices [16]. A strength of this work is the number of responses to the survey. Previous research has obtained relatively small numbers of peer supporters discussing a particular programme or intervention whereas this is more about getting insights into the wider picture.

Peer support programs often involve recruiting and training peer supporters from local communities. This not only creates employment opportunities but also fosters a sense of community empowerment and cohesion. Local peer supporters can better understand the unique challenges and needs of their communities, making the support they provide more relevant and impactful. Furthermore, experienced peer supporters often have a good idea of what is going on in their local areas and can help to identify gaps and specific needs. This study highlights some gaps in potential training and infrastructure support (e.g. funding).

It must be noted that the majority of peer supporters responding to this survey were active as part of a breastfeeding organisation rather than within the NHS. Breastfeeding peer support programs are often integrated into existing healthcare services, such as maternity care or public health initiatives [9]. This aims to ensure that peer support is accessible to all mothers and complements the support provided by healthcare professionals. However, it has previously been noted that breastfeeding peer support services are not well integrated with NHS health services [9]. Future efforts are needed to integrate breastfeeding peer support services within the NHS in Wales.

The UK has one of the lowest breastfeeding rates in the world. The last Infant Feeding Survey (IFS) was conducted in 2010 [17] and reported that though 80% of babies are breastfed at birth, only 1% are exclusively breastfed for 6 months in the UK [17]. In 2022, 61.7% of babies in Wales were breastfed at birth, which dropped to 19.3% being exclusively breastfed at 6 months [18]. It must be highlighted that the IFS and Wales statistics used different measures of exclusive breastfeeding. Moreover, it is important to note that with the reduction in the number of women choosing to breastfeed, there will be a reduction in the pool of future peer supporters. Therefore, peer support is just one tool we have and needs to be considered as part of a holistic strategy to improve breastfeeding rates. Peer supporters provide a supportive community where breastfeeding is normalised despite low prevalence. Future research could aim to explore how we retain those who do currently give up their time to volunteer as breastfeeding peer supporters. Perhaps a national policy or guidance needs to consider business continuity from a workforce perspective.

Despite efforts to make peer support accessible, there may still be challenges in reaching all mothers, particularly those in rural or marginalised communities [9]. Previous research has highlighted that breastfeeding peer support is not always accessed by mothers living in more deprived areas [9]. Limited availability of trained peer supporters, as well as logistical barriers such as transportation or language barriers, can hinder access to support for some mothers who may need it the most.

This survey did not ask any questions relating to volunteer demographics. This is a potential weakness as factors that may influence the decision to provide breastfeeding peer support (e.g. age, ethnicity, socioeconomic status etc) cannot be explored. The large number of responses to this survey was not anticipated. It was felt demographic data could be potentially identifying and would not be representative in the small numbers anticipated. Follow-up exploration of the demographics of those who undertake breastfeeding peer support training may reveal insights. While peer support programs aim to be inclusive, there may still be equity considerations to address. Certain groups of mothers, such as those from low-income backgrounds or minority ethnic groups, may face additional barriers to accessing peer support.

Specific shared cultural experience with peer supporters can help mothers to overcome specific barriers [19]. Self funding for peer support services may also have an impact on equity. Efforts to address these disparities and ensure equitable access to support are essential for maximising the impact of breastfeeding peer support programs in Wales.

## Conclusions

Breastfeeding peer support programs in Wales play a valuable role in promoting and supporting breastfeeding among mothers. While volunteers reported positive experiences in fostering community empowerment, challenges remain in ensuring accessibility, quality assurance, sustainability, and equity. Addressing these challenges requires continued investment, collaboration between stakeholders, and a commitment to providing comprehensive and culturally sensitive support to all mothers in Wales. Breastfeeding peer support volunteers play a significant role for new breastfeeding mothers and these respondents provided passionate responses regarding their experiences. The views expressed will directly inform work carried out under the AWBAP [12]. Overall, breastfeeding peer support programs have the potential to make a significant contribution to improving maternal and child health outcomes in Wales, but ongoing efforts are needed to maximise their effectiveness and reach.

## Data Availability

All data produced in the present study are available upon reasonable request to the authors

