## Supplementary Information for "Experiences of breastfeeding peer support: views from current, past and trainee breastfeeding peer support volunteers in Wales, UK"

| **Survey Question** | **Response Options** |
| --- | --- |
| Are you currently active as a peer support volunteer? | - No, trained/training but never/not yet been active - Yes, with local NHS - Yes, with a breastfeeding organisation - Previously active in NHS - Previously active with breastfeeding organisation |
| With which organisation/s did you complete peer support training? | - Association of Breastfeeding Mothers (ABM) - Breastfeeding Network (BfN) - National Childbirth Trust (NCT) - Local NHS - Agored - Other |
| How long ago did you complete your training? | - Less than 12 months - 1-2 years - 2-3 years - 3-5 years - 5 years plus - I am training now |
| How long have you been/were you active as a volunteer peer supporter? | - Up to six months - Six to twelve months - 12-24 months - 2-4 years - 4 years plus (please specify in “other”) - Trained in past but did not actively volunteer - Training and not yet active - Other |
| Why did you decide to undertake peer support training? |  |
| In which health board area do/have you volunteered? | - Anuerin Bevan University Health Board - Betsi Cadwaladr University Health Board - Cardiff and Vale University Health Board - Cwm Taf Morgannwg University Health Board - Hywel Dda University Health Board - Powys Teaching Health Board - Swansea Bay University Health Board - Outside Wales - Prefer not to say - Other |
| Training costs | - Self funded - NHS funded - Charity project funded - Not applicable |
| Childcare during training | - Self funded  - NHS funded  - Charity project funded  - Not applicable |
| Expenses incurred during training | - Self funded  - NHS funded  - Charity project funded  - Not applicable |
| DBS check costs. | - Self funded  - NHS funded  - Charity project funded  - Not applicable |
| Expenses incurred during volunteering | - Self funded  - NHS funded  - Charity project funded  - Not applicable |
| Have you volunteered in any other capacity related to infant feeding, e.g. breastfeeding counsellor? If so, please tell us more. |  |
| Please select the relevant option to go to the next section: | - Currently active volunteers - Previously active volunteers - Never/not yet active volunteers |
| **Active volunteers** | |
| Which of these activities do you take part in as a volunteer? Please choose all that apply. | - Support mothers in the hospital setting - Attend a community group with healthcare professionals in attendance - Attend a community group with a breastfeeding counsellor/IBCLC - Lead a community group with other peer supporters - Provide support on social media e.g. Facebook group - Conduct home visits - Take part in antenatal classes/groups - Take part in a specific peer support intervention for research purposes - Lead a community group alone - Other |
| How long do you currently spend doing volunteer peer support each week (hours)? | - Up to 2 hours - 2-4 hours - Over 4 hours – please specify in “other” - Other |
| What is your experience of volunteering with your local health board? | - Prefer to volunteer outside of NHS - No NHS opportunities available - Looked into it but barriers put me off - I am an NHS volunteer and found the process difficult - I am an NHS volunteer and found the process easy - Other |
| I have a clear understanding of the role and boundaries of a peer supporter. | - Yes - No - Not applicable |
| I have access to regular supervision. | - Yes - No - Not applicable |
| I have access to regular update training. | - Yes - No - Not applicable |
| I know who to contact for problems which are out of my scope and how to get hold of them. | - Yes - No - Not applicable |
| I have undertaken NHS volunteer training/ induction. | - Yes - No - Not applicable |
| I have a current DBS check. | - Yes - No - Not applicable |
| Thinking about your experience of peer support over the last six months, can you tell us about the most common issues you have been asked for support with? |  |
| **Previously active volunteers** | |
| Which of these activities did you take part in as a volunteer? Please choose all that apply. | - Support mothers in the hospital setting - Attend a community group with healthcare professionals in attendance - Attend a community group with a breastfeeding counsellor/IBCLC - Lead a community group with other peer supporters - Provide support on social media e.g. Facebook group - Conduct home visits - Take part in antenatal classes/groups - Take part in a specific peer support intervention for research purposes - Lead a community group alone - Other |
| Why did you stop volunteering as a peer supporter? (Please choose all that apply). | - Too busy to continue - Returned to work post maternity leave - Project/opportunity ended - Covid pandemic - Moved onto another breastfeeding related role or training e.g. breastfeeding counsellor, International Board Certified Lactation Consultant - Started midwifery or nursing training - Started a paid role related to infant feeding - Other |
| **All peer supporters** | |
| Which, if any, of the following do you think will be useful to breastfeeding peer supporters and infant feeding teams in Wales in the future? | |
| A national job description for NHS peer support volunteers | - Essential - Useful - Nice to have - Not useful ay all - No opinion |
| National guidance for infant feeding teams about peer support in the NHS | - Essential - Useful - Nice to have - Not useful ay all - No opinion |
| A list of recognised training organisations | - Essential - Useful - Nice to have - Not useful ay all - No opinion |
| National volunteer awards | - Essential - Useful - Nice to have - Not useful ay all - No opinion |
| National guidance for health board volunteer services | - Essential - Useful - Nice to have - Not useful ay all - No opinion |
| National peer support training/ qualification | - Essential - Useful - Nice to have - Not useful ay all - No opinion |
| National volunteer coordinator | - Essential - Useful - Nice to have - Not useful ay all - No opinion |
| Can you choose three words to describe your experience of breastfeeding peer support? |  |
| Please use this box to share anything you would like to about your experience of volunteering as a peer supporter and/or your thoughts about the work we are doing. |  |
